# Exploring the Role of Superspreading Events in SARS-CoV-2 Outbreaks

**DOI:** 10.1101/2022.06.29.22277010

**Authors:** Jordan Bramble, Alexander Fulk, Raul Saenz, Folashade B. Agusto

## Abstract

The novel coronavirus SARS-CoV-2 emerged in 2019 and subsequently spread throughout the world, causing over 529 million cases and 6 million deaths thus far. In this study, we formulate a continuous-time Markov chain model to investigate the influence of superspreading events (SSEs), defined here as public or social events that result in multiple infections over a short time span, on SARS-CoV-2 outbreak dynamics. Using Gillespie’s direct algorithm, we simulate a continuous-time Markov chain model for SARS-CoV-2 spread under multiple scenarios: first, with neither hospitalisation nor quarantine; second, with hospitalisation, quarantine, premature hospital discharge, and quarantine violation; and third, with hospitalisation and quarantine but neither premature hospital discharge nor quarantine violation. We also vary quarantine violation rates. Results indicate that, in most cases, SSE-dominated outbreaks are more variable but less severe than non-SSE-dominated outbreaks, though the most severe SSE-dominated outbreaks are more severe than the most severe non-SSE-dominated outbreaks. SSE-dominated outbreaks are outbreaks with relatively higher SSE rates. In all cases, SSE-dominated outbreaks are more sensitive to control measures, with premature hospital discharge and quarantine violation substantially reducing control measure effectiveness.

## 1 Introduction

Severe acute respiratory syndrome coronavirus 2 (SARS-CoV-2) is the causative agent of COVID-19. Since emerging in China’s Hubei province in 2019, it has caused over 529 million cases and 6 million deaths worldwide [47], with over 84 million cases and 1 million deaths in the United States [16]. With an estimated basic reproduction number (R_0_) ranging from 2 to 4 [13], the virus is moderately infectious. COVID-19 has strained the U.S. healthcare system, with many hospitals nearing or exceeding capacity [2, 34, 24, 19] and some moving to ration care [8, 9, 11, 17, 25]. In response, both national and state-level governments have issued guidelines and mandates aimed at reducing transmission, ranging from social-distancing guidelines and mask mandates to stay-at-home orders and limits on large gatherings [36, 40].

The effectiveness of these guidelines and mandates has been hindered by imperfect adherence and compliance. For example, some people refuse to wear a mask [21]. Moreover, many people fail to social distance, despite doing so initially [23]. People also violate stay-at-home orders, with several being issued citations and some being arrested [22, 29, 31, 45]; more people than indicated by citations and arrests alone have likely violated stay-at-home orders, given the variability in enforcement protocols [4, 18]. Beyond refusing to mask, failing to social distance, and violating stay-at-home orders, some people also attend public or social events. This facilitates superspreading events (SSEs), defined here as public or social events that result in multiple infections over a short time span. Such events have contributed to SARS-CoV-2 spread [30]. All of the aforementioned are functions of human behavior.

SSEs differ from superspreading individuals (SIs), which we define here as individuals who cause disproportionately more infections over their infectious lifetime. Event- and individual-based superspreading are not mutually exclusive; people who cause SSEs may qualify as SIs. However, this is not always the case. People may cause multiple infections at a public/social event but not cause disproportionately more infections over their infectious lifetime. Likewise, not all who become SIs do so by causing SSEs. People may become SIs due to intrinsic factors, such as greater-than-average contact rates or viral shedding [3, 32]. Heterogeneity in contact rates is well-established, while heterogeneity in SARS-CoV-2 shedding is evident in Badu, et al’s literature review [7] on SARS-CoV-2 viral loads, shedding, and transmission dynamics. Only a subset of the population may thus achieve SI status without causing SSEs, whereas anyone may cause SSEs under the right circumstances. Extrinsic factors such as crowding and poor ventilation potentiate SSEs [3, 6].

Event- and individual-based superspreading are incorporated into models using different frameworks. SSEs may be modeled via rare events resulting in multiple infections; these events may be caused by any individual, and their frequency and number of resulting infections each follow some distribution. This is the approach taken by James, et al [26]. SIs may be modeled via heterogeneity in infectivity; individual infectivity follows some distribution, the right tail of which corresponds to superspreader individuals. This is the approach taken by Lloyd, et al [32]. There are many other approaches to modeling SIs [43, 37, 42, 38]. Note that while some of these articles mention superspreading events, their frameworks are nonetheless individual-based. Whereas most superspreading-incorporating models use an individual-based framework, we use an events-based framework.

In what follows, SSE-dominated outbreaks refer to outbreaks with relatively higher SSE rates than non-SSE-dominated outbreaks; non-SSEs refer to non-SSE-related infection events. The goals of this study are to investigate:

G1. The influence of SSEs relative to that of non-SSEs on outbreak dynamics

G2. The effectiveness of hospitalisation and quarantine as control measures for SSE- versus non-SSE-dominated outbreaks

G3. The influence of quarantine violation on the effectiveness of quarantine for SSE- versus non-SSE-dominated outbreaks

We incorporate SSEs into a continuous-time Markov chain (CTMC) model, impose a constancy condition, and vary SSE and non-SSE rates to accomplish G1. The constancy condition requires that the expected number of infections following the CTMC model’s first change in state remain constant for different SSE and non-SSE rates. We simulate the CTMC model under multiple scenarios to accomplish G2:

(i) With neither hospitalisation nor quarantine (NHQ). This scenario excludes hospitalization and quarantine.

(ii) With realistic hospitalization and quarantine (RHQ). This scenario includes hospitalisation, quarantine, premature hospital discharge, and quarantine violation.

(iii) With idealistic hospitalization and quarantine (IHQ). This scenario includes hospitalisation and quarantine but excludes premature hospital discharge and quarantine violation.

While simulating the CTMC model under these different scenarios partially addresses G3, we also simulate it under RHQ with varying levels of quarantine violation to accomplish G3. Our methods, results, and discussion are located in Sections 2, 3, and 4, respectively.

## 2 Methods

We derive our continuous-time Markov chain (CTMC) model from Agusto et. al.’s [1] baseline COVID-19 model (see Section 2.1) and incorporate SSEs in Section 2.2. The constancy condition is derived in Section 2.3. After formulating the CTMC model and imposing the constancy condition, we detail our simulation protocol in Section 2.4. Finally, we introduce two superspreading-incorporating, discrete-time Markov chain (DTMC) models from literature [32, 26] in Section 2.5. These models’ results are used for comparison in Section 4.

### 2.1 Baseline COVID-19 Model

In Agusto et al’s baseline COVID-19 model [1], the evolution of susceptible (*S*(*t*)), exposed (*E*(*t*)), asymptomatic (*A*(*t*)), symptomatic (*I*(*t*)), hospitalized (*H*(*t*)), quarantined (*Q*(*t*)), and removed (*R*(*t*)) individuals through time is governed by the system of ordinary differential equations given below:

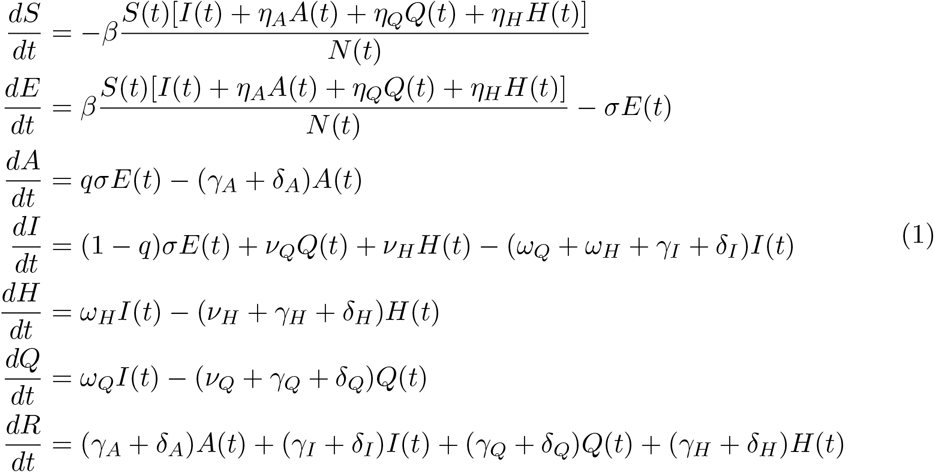

The exposed and asymptomatic classes account for the virus’ incubation period and the reduced infectiousness of asymptomatic individuals. Meanwhile, hospitalisation and quarantine function as control measures. Movement of individuals from the hospitalized and quarantined classes back to the symptomatic class account for limited resources and human behavior. We refer to the former as premature hospital discharge and the latter as quarantine violation.

The model’s flow diagram is displayed in Figure 1, and its parameters are defined in Table 1.

**Figure 1:**
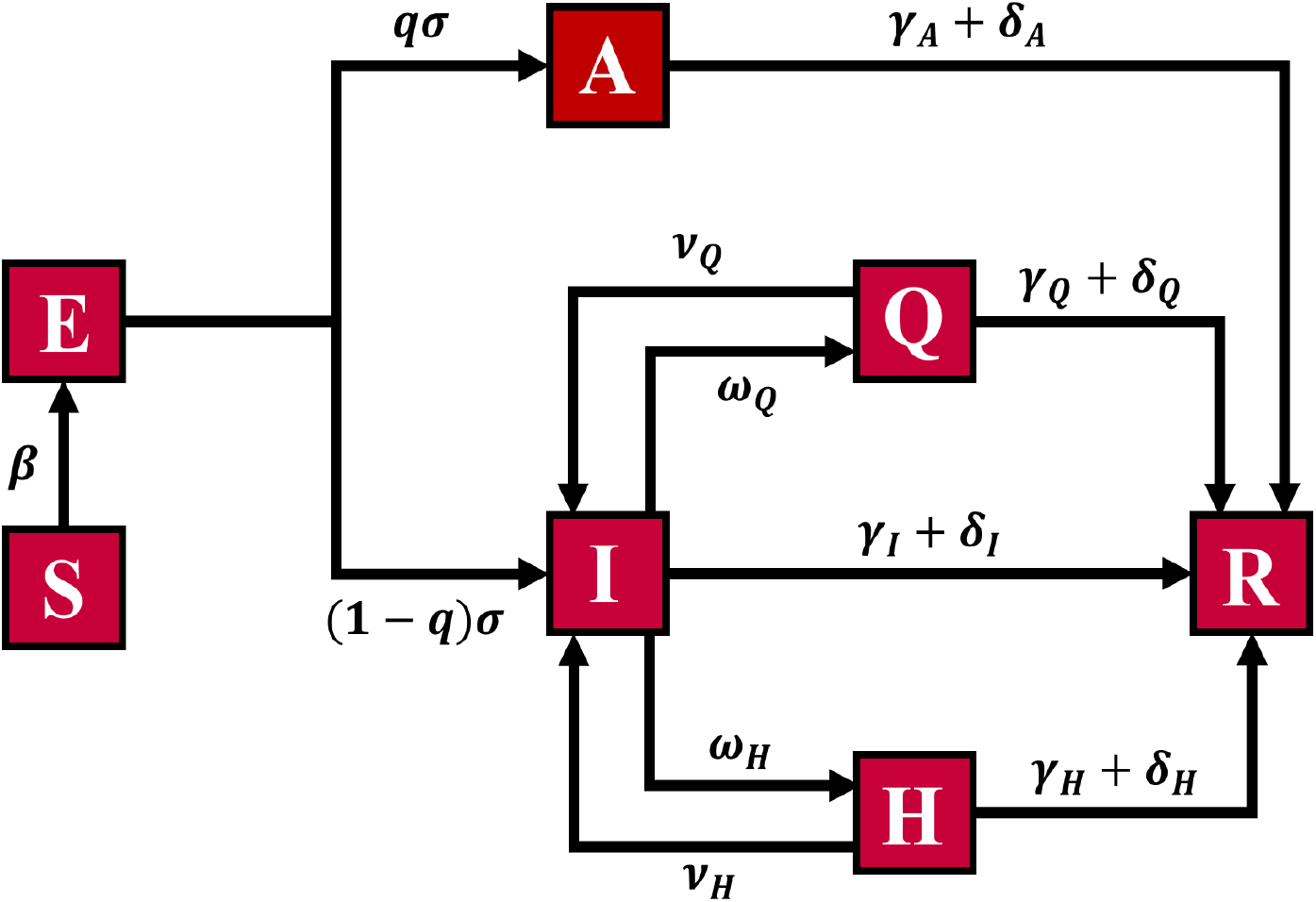
Compartmental flow diagram for the baseline COVID-19 model (1). Susceptible individuals are exposed upon initial infection, and exposed individuals are either asymptomatic or symptomatic once infectious. Asymptomatic individuals are removed upon recovery or death, while symptomatic individuals may be hospitalised or quarantined and are removed upon recovery or death. Hospitalised and quarantined individuals may be prematurely discharged from the hospital or violate quarantine and are removed upon recovery or death.

**Table 1:** Parameter values for the baseline COVID-19 model (1) from Agusto et. al. [1].

| Parameter | Description | Value |
| --- | --- | --- |
| $\beta$ | Infection rate | 0.4975 |
| $\eta_A, \eta_H, \eta_Q$ | Infection rate modifiers | 0.45, 0.1362, 0.3408 |
| $q$ | Proportion that remain asymptomatic | 0.5 |
| $\sigma$ | Disease progression rate | $\frac{1}{6}$ |
| $\gamma_A, \gamma_I, \gamma_H, \gamma_Q$ | Recovery rates | 0.7565, 0.0775, 0.041, 0.083 |
| $\omega_H, \omega_Q$ | Hospitalization & quarantine rates | 0.1977, 0.453 |
| $\nu_H, \nu_Q$ | Hospital discharge & quarantine violation rates | 0.1301, 0.4605 |
| $\delta_A, \delta_I, \delta_H, \delta_Q$ | Death rates | 0.00325, 0.0065, 0.0065, 0.0065 |

### 2.2 Continuous-Time Markov Chain Model

We limit ourselves to outbreaks scenarios – specifically, the short time period following the introduction of a small number of infected individuals into a completely susceptible population – so that we may assume the following:

(†) Population size and mixing is such that transmission events are independent of each other and unaffected by the depletion of susceptible individuals and accumulation of removed individuals [35].

This assumption is reasonable in outbreak scenarios because the number of infected individuals is small relative to the number of susceptible individuals. (†) implies that 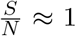, so we may approximate the rate at which individuals transition from *S* to *E* due to non-SSEs as

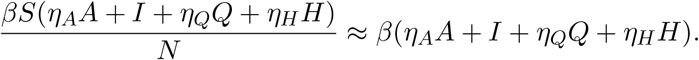

The possible events corresponding to the baseline COVID-19 model (1) are:

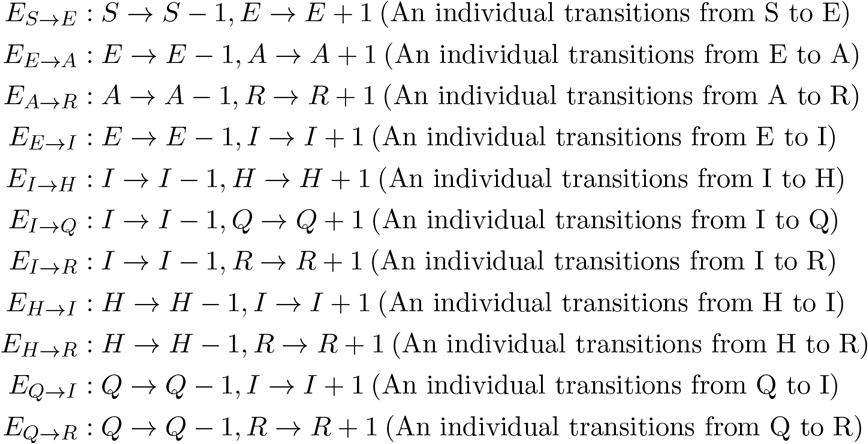

*E*_*S→E*_ corresponds to a susceptible individual being exposed; *E*_*E→A*_ corresponds to an exposed individual becoming infectious but remaining asymptomatic; *E*_*A→R*_ corresponds to an asymptomatic individual recovering or dying; *E*_*E→I*_ corresponds to an exposed individual becoming infectious and symptomatic; *E*_*I→H*_ corresponds to a symptomatic individual being hospitalised; *E*_*I→Q*_ corresponds to a symptomatic individual being quarantined; *E*_*I→R*_ corresponds to a symptomatic individual recovering or dying; ; *E*_*H→I*_ corresponds to a hospitalised individual being prematurely discharged from the hospital; and *E*_*H→R*_ corresponds to a hospitalised individual recovering or dying; *E*_*Q→I*_ corresponds to a quarantined individual violating quarantine; *E*_*Q→R*_ corresponds to a quarantined individual recovering or dying.

Let *K* ∈ ℕ ∪{0} be a Poisson random variable with expectation *ϕ* ∈ ℕ. We incorporate SSEs by defining an additional event,

*E*_*SSE*_ : *S* → *S* − *k, E* → *E* + *k* (*k* individuals transition from S to E), where *k* is some possible value of *K. E*_*SSE*_ corresponds to *k* susceptible individuals becoming exposed. The event thus generates a random number of infections over a short period of time. This differs from *E*_*S→E*_, which generates one infection. In what follows, we take *ψ* ∈ ℝ^+^ to be the deterministic rate at which infected individuals cause SSEs. (†) implies that SSEs involve a single infected individual and otherwise susceptible individuals, and assuming that hospitalized and quarantined individuals do not cause SSEs, we may take *ψ*(*η*_*A*_*A* + *I*) to be the deterministic rate at which SSEs occur.

Now, let *τ* ∈ ℝ^+^. By considering the above events’ occurrence in a sufficiently small time interval, (*τ, τ* +Δ*τ*), we may assume that at most one event occurs in this interval. The probabilities corresponding to each event’s occurrence in (*τ, τ* + Δ*τ*) are then:

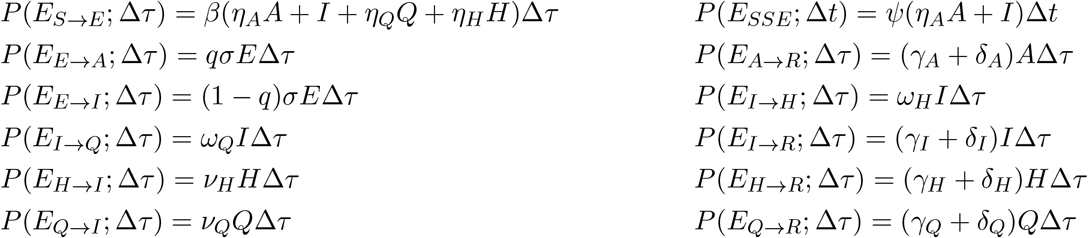

Δ*τ* is taken to be small enough that Σ*P* (*E*_*ξ*_; Δ*τ*) ≤ 1 for all *τ* ≤ *T*, where *ξ* ∈ {*S* → *E, SSE, E* → *A, A* → *R, E* → *I, I* → *H, I* → *Q, I* → *R, H* → *I, H* → *R, Q* → *I, Q* → *R*} and *T* ∈ ℝ^+^; this ensures that the above are valid probabilities. We denote the set of all possible transitions as 𝕊_*T*_. The stochastic instantaneous rates at which events occur are obtained by dividing the above probabilities by Δ*τ* and taking

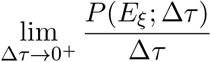

for each *ξ*:

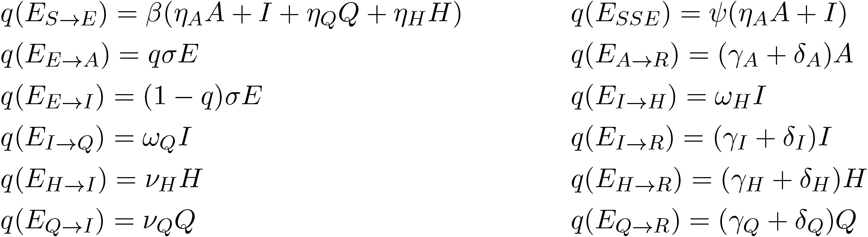

From the stochastic instantaneous rates, we obtain the probabilities of given events being the next to occur. Letting Ω ≐ ∑ *q*(*E*_*ξ*_), we have:

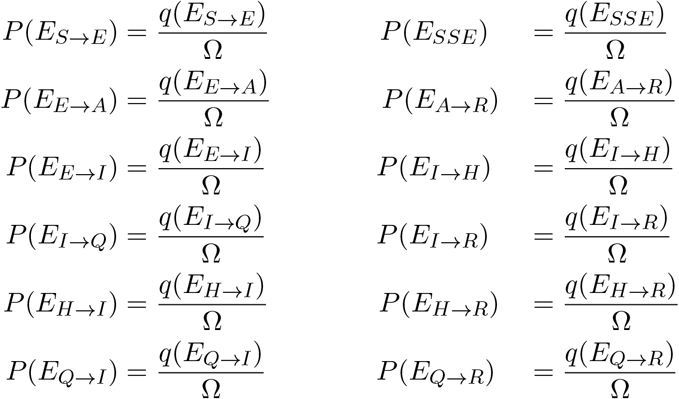

The set of events 𝕊_*E*_ ≐ {*E*_*ξ*_ : *ξ* ∈ 𝕊_*T*_ } and probabilities 𝕊_*P*_ ≐ {*P* (*E*_*ξ*_) : *ξ* ∈ 𝕊_*T*_ } constitute a continuous-time Markov chain. Our CTMC derivation from the baseline COVID-19 model (1) is adapted from Oluwatobilloba’s CTMC derivations from simpler (SIS and SIR) models for infectious disease spread [39]. For further reading on infectious disease modeling, see [28], and for general reading on Markov chains, see [41, 10].

### 2.3 Constancy Condition

To investigate the relative influence of SSEs versus non-SSEs on outbreak dynamics, we impose a constancy condition by requiring that the expected number of infections following the first change in state remain constant for different rates of SSEs and non-SSEs. We denote this expectation as 𝔼_0_(*X*), where *X* ∈ ℕ ∪ {0} is a random variable corresponding to the number of infections following a change in state. Note that these infections may be SSE- or non-SSE-related. Our constancy condition is adapted from James et al’s [26] constancy condition. Assuming that James et al take *I*_0_ = 1, both conditions may be derived using the pgfs of *X* given the systems’ initial states. However, the interpretation of *E*_0_(*X*) differs, as the systems’ state changes have different meanings (see Section 2.5).

Let *β*^*∗*^ = *κβ*, where *κ* ∈ [0, 1] and *β* is the fitted infection rate parameter (see Table 1), which we take to be the non-SSE rate in the absence of SSEs. This gives *β*^*∗*^ ∈ [0, *β*]. We seek *ψ*, the SSE rate, such that the constancy condition is satisfied for an arbitrary *β*^*∗*^. To begin, we derive the pgf for *X*. Letting

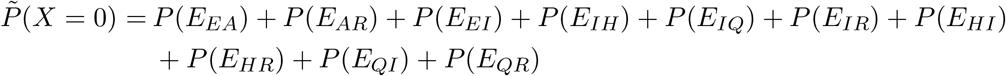

and recalling that *K* ∼ Poisson(*ϕ*), we have:

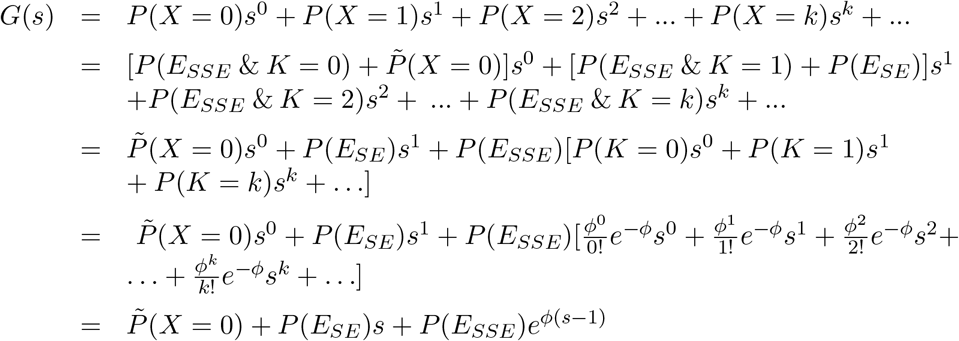

From the pgf, we obtain the expectation of *X*:

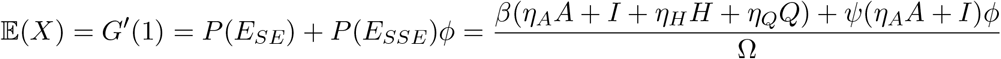

Taking ⟨*E, A, I, H, Q*⟩ = ⟨*E*_0_, *A*_0_, *I*_0_, *H*_0_, *Q*_0_⟩, where the latter are the initial numbers of exposed, asymptomatic, symptomatic, hospitalised, and quarantined individuals, 𝔼(*X*) becomes 𝔼_0_(*X*). For further reading on pgfs and their properties and epidemiological applications, see [35, 44].

Next, we obtain *ψ* as a function of *β*^*∗*^. Letting 𝔼_0,*β*_(*X*) and 𝔼_0,*β*_*∗* (*X*) be the initial expectations when the non-SSE rates are *β* and *β*^*∗*^ *< β* and setting *E*_0,*β*_(*X*) = *E*_0,*β*_*∗* (*X*),we have:

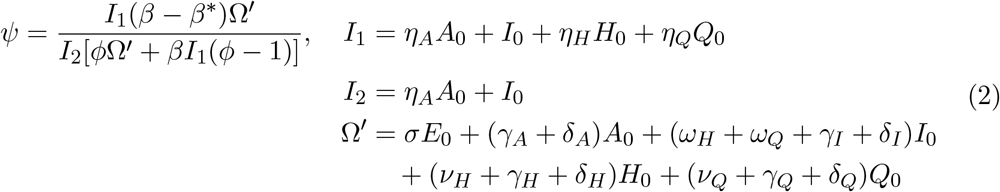

Note that *ψ* is only defined for *I*_2_, *ϕ*Ω^′^ + *βI*_1_(*ϕ* − 1) > 0. The requirement that *ψ* be of the above form constitutes our constancy condition.

Also note that *ψ* is invariant under scaling with respect to ⟨*E*_0_, *A*_0_, *I*_0_, *H*_0_, *Q*_0_⟩. Indeed, letting 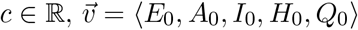, and 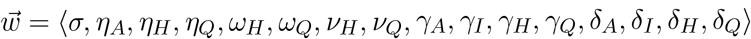 and noting that, and 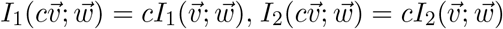, and 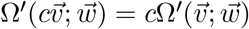 we have:

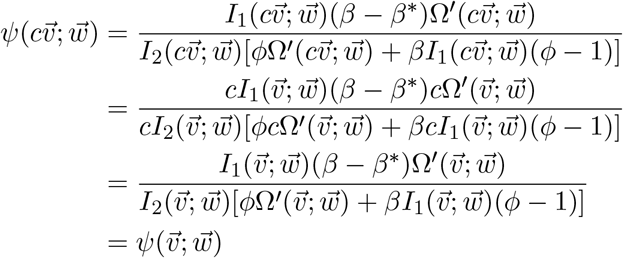

This property motivates our choice of initial conditions when simulating the CTMC model (see Section 2.4).

### 2.4 Model Simulation

Because our model is a CTMC, it may be simulated using Gillespie’s direct algorithm [20, 28]. The algorithm is a direct (versus approximate) method for simulating stochastic processes [20]. We simulated the model under the following scenarios:

i. Neither hospitalization nor quarantine (NHQ), in which hospitalisation, quarantine, premature hospital discharge, and quarantine violation are excluded from the model; this is equivalent to excluding *E*_*I→H*_, *E*_*I→Q*_, *E*_*H→I*_, and *E*_*Q→I*_ from the model
ii. Realistic hospitalization and quarantine (RHQ), in which hospitalisation, quarantine, premature hospital discharge, and quarantine violation are included in the model; this is equivalent to including *E*_*I→H*_, *E*_*I→Q*_, *E*_*H→I*_, and *E*_*Q→I*_ in the model
iii. Idealistic hospitalization and quarantine (IHQ), in which hospitalisation and quarantine are included in the model but premature hospital discharge and quarantine violation are excluded; this is equivalent to including *E*_*I→H*_ and *E*_*I→Q*_ in the model but excluding *E*_*H→I*_ and *E*_*Q→I*_

Note that RHQ corresponds to the baseline COVID-19 model (1). We also simulated the model under RHQ with low quarantine violation (*lqv*) and high quarantine violation (*hqv*). We denote these sub-scenarios as RHQ_*lqv*_ and RHQ_*hqv*_. For *lqv*, we halved the fitted value of quarantine violation 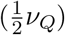, and for *hqv*, we doubled the fitted value of quarantine violation (2*ν*_*Q*_).

Events were excluded by setting their corresponding stochastic instantaneous rates to zero. In NHQ, *ω*_*H*_, *ω*_*Q*_, *ν*_*H*_, and *ν*_*Q*_ were set to zero, thereby eliminating hospitalization, quarantine, premature hospital discharge, and quarantine violation; in RHQ, *ω*_*H*_, *ω*_*Q*_, *ν*_*H*_, and *ν*_*Q*_ remained set to their fitted values (see Table 1); and in IHQ, *ω*_*H*_ and *ω*_*Q*_ remained set to their fitted values, while *ν*_*H*_ and *ν*_*Q*_ were set to zero, thereby eliminating premature hospital discharge and quarantine violation.

The initial conditions for NHQ, RHQ, and IHQ were ⟨*E*_0_, *A*_0_, *I*_0_, *H*_0_, *Q*_0_⟩ = ⟨15, 2, 4, 0, 0⟩, ⟨14, 2, 2, 1, 1⟩, and ⟨14, 2, 2, 1, 1⟩, respectively; the CTMC model is independent of *S*_0_ and *R*_0_. These initial conditions were obtained by simulating the baseline COVID-19 model (1) under each scenario with ⟨*S*_0_, *E*_0_, *A*_0_, *I*_0_, *H*_0_, *Q*_0_, *R*_0_⟩ = ⟨10^10^, 20, 0, 0, 0, 0, 0⟩ and recording the number of individuals in each class on day 3. *S*_0_ was taken to be 10^10^ to ensure that the depletion of susceptible individuals had a negligible influence. Recall that the constancy condition is not defined for 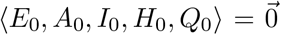 and is invariant under scaling of ⟨*E*_0_, *A*_0_, *I*_0_, *H*_0_, *Q*_0_⟩ (see Section 2.3). This motivates the need for a realistic initial proportion of asymptomatic, symptomatic, hospitalised, and quarantined individuals. The initial conditions for RHQ_*lqv*_ and RHQ_*lqv*_ were taken to be the same as for RHQ.

For NHQ, RHQ, and IHQ, *κ* was varied from 0 to 1 in increments of 0.1, and *ψ* was calculated for each *β*^*∗*^ value using the constancy condition (2) with scenarios’ corresponding initial conditions and parameter sets. *κ* was also varied from 0 to 1 in increments of 0.1 for RHQ_*lqv*_ and RHQ_*hqv*_, but *ψ* was taken to be the same as for RHQ; it was not re-calculated using 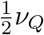 or 2*ν*_*Q*_. This allows for better isolation of quarantine violation’s influence on quarantine effectiveness for SSE- versus non-SSE- dominated outbreaks.

Simulations were ended once either the disease went extinct or 50 active infections were attained, similar to the approach taken in [32]. The number of extinctions and total simulations were recorded to estimate the probabilities of outbreak extinction, and for surviving outbreaks, the times at which 50 active infections were attained and the cumulative numbers of SSE-related and non-SSE-related infections were recorded for analysis. These times are hereafter referred to as stop times.

The number of simulations depended on the variances of the stop times. If the variance was less than 1500, 50000 simulations were ran; if the variance was between 1500 and 15000, 500000 simulations were ran; and if the variance was greater than 15000, 1500000 simulations were ran. Table 2 gives the number of simulations for each parameter set.

**Table 2:**
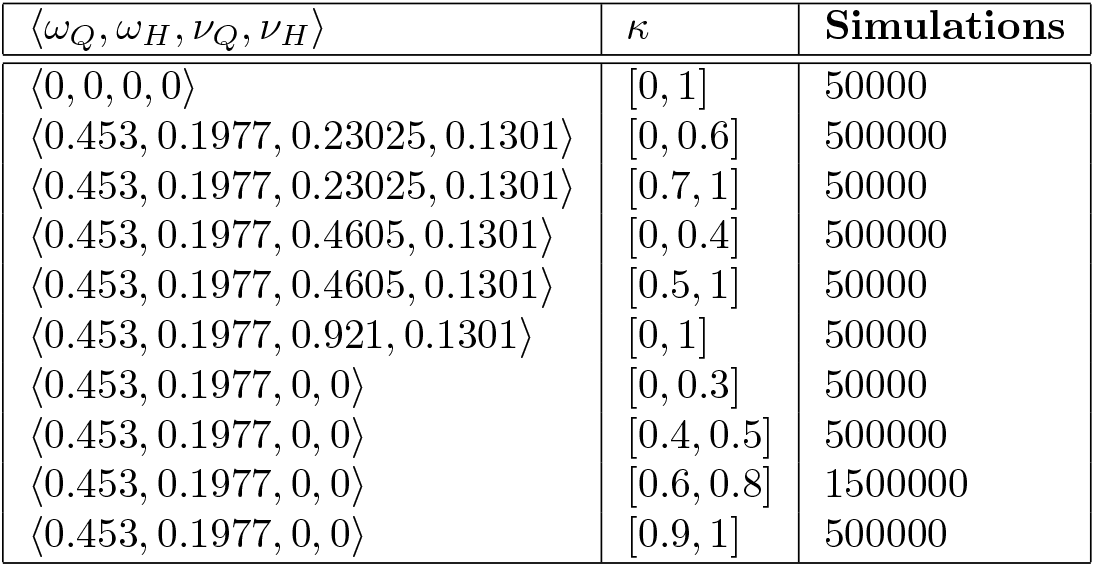
Number of simulations for each parameter set. ⟨0, 0, 0⟩ corresponds to NHQ, ⟨0.453, 0.1977, 0.23025, 0.1301⟩ corresponds to RHQ_*lqv*_, ⟨0.453, 0.1977, 0.4605, 0.1301⟩ corresponds to RHQ, ⟨0.453, 0.1977, 0.921, 0.1301⟩ corresponds to RHQ_*hqv*_, and ⟨0.453, 0.1977, 0, 0⟩ corresponds to IHQ.

| $\langle \omega_Q, \omega_H, \nu_Q, \nu_H \rangle$ | $\kappa$ | Simulations |
| --- | --- | --- |
| $\langle 0, 0, 0, 0 \rangle$ | $[0, 1]$ | 50000 |
| $\langle 0.453, 0.1977, 0.23025, 0.1301 \rangle$ | $[0, 0.6]$ | 500000 |
| $\langle 0.453, 0.1977, 0.23025, 0.1301 \rangle$ | $[0.7, 1]$ | 50000 |
| $\langle 0.453, 0.1977, 0.4605, 0.1301 \rangle$ | $[0, 0.4]$ | 500000 |
| $\langle 0.453, 0.1977, 0.4605, 0.1301 \rangle$ | $[0.5, 1]$ | 50000 |
| $\langle 0.453, 0.1977, 0.921, 0.1301 \rangle$ | $[0, 1]$ | 50000 |
| $\langle 0.453, 0.1977, 0, 0 \rangle$ | $[0, 0.3]$ | 50000 |
| $\langle 0.453, 0.1977, 0, 0 \rangle$ | $[0.4, 0.5]$ | 500000 |
| $\langle 0.453, 0.1977, 0, 0 \rangle$ | $[0.6, 0.8]$ | 1500000 |
| $\langle 0.453, 0.1977, 0, 0 \rangle$ | $[0.9, 1]$ | 500000 |

### 2.5 Discrete-Time Markov Chain Models

With CTMC models, the system may assume a new state at any time *t* ∈ ℝ^+^; *t* is thus continuous. For our CTMC model, every time-step corresponds to a single event *E*_*ξ*_, *ξ* ∈ 𝕊_*T*_, occurring. With discrete-time Markov chain (DTMC) models, the system may only assume a new state at times {*t*_*k*_ : *k* ∈ ℕ }; *t* is thus discrete. For Lloyd et al’s [32] and James et al’s [26] DTMC models, every time-step corresponds to the death of the current generation of infected individuals and birth of the next generation of infected individuals. The CTMC and DTMC models’ state changes thus have different meanings.

Lloyd, et al [32] utilize an individual-based superspreading framework. They incorporate SIs into their DTMC via the random variable *ν*, the expected number of infections caused by an individual. Individuals thus have different potentials to infect others. They simulate their model under multiple scenarios, each of which assumes a different distribution for *ν*. We limit our comparison (see Section 4) to the scenario which assumes *ν* is *γ*-distributed. James, et. al. [26] utilize an event-based framework. They incorporate SSEs into their DTMC via *ρ*, the expected number of SSEs caused by an individual, and *λ*, the expected number of infections caused by an SSE. Every individual has the same potential to infect others.

## 3 Results

In each figure, *κ* increases from 0 to 1 in increments of 0.1; increasing values of *κ* correspond to decreasing rates of SSEs and increasing rates of non-SSEs. Note that, while all figures’ x-axes are the same, their y-axes differ.

### Scenario (i): Neither Hospitalisation nor Quarantine (NHQ)

In NHQ, hospitalization, quarantine, premature hospital discharge, and quarantine violation are excluded from the model. Figure 2(a) shows the distribution of stop times. The maximum, mean, and median stop times strictly decrease for *κ* ∈ [0, 1], while the minimum stop times are similar for *κ* ∈ [0, 0.7) but strictly increase for *κ* ∈ [0.7, 1]. Figures 2(b) and 2(c) show the variances of stop times and probabilities of extinction, respectively. Both strictly decrease for *κ* ∈ [0, 1], but they do so more quickly for smaller *κ*. Figure 2(d) shows the means of the cumulative numbers of SSE- related and non-SSE-related infections. Their curves are labelled as SSE and Non-SSE in the legend. The SSE curve strictly decreases with increasing *κ*, while the Non-SSE curve strictly increases. They do so more quickly for smaller *κ* and intersect between *κ* = 0.4 and *κ* = 0.5.

**Figure 2:**
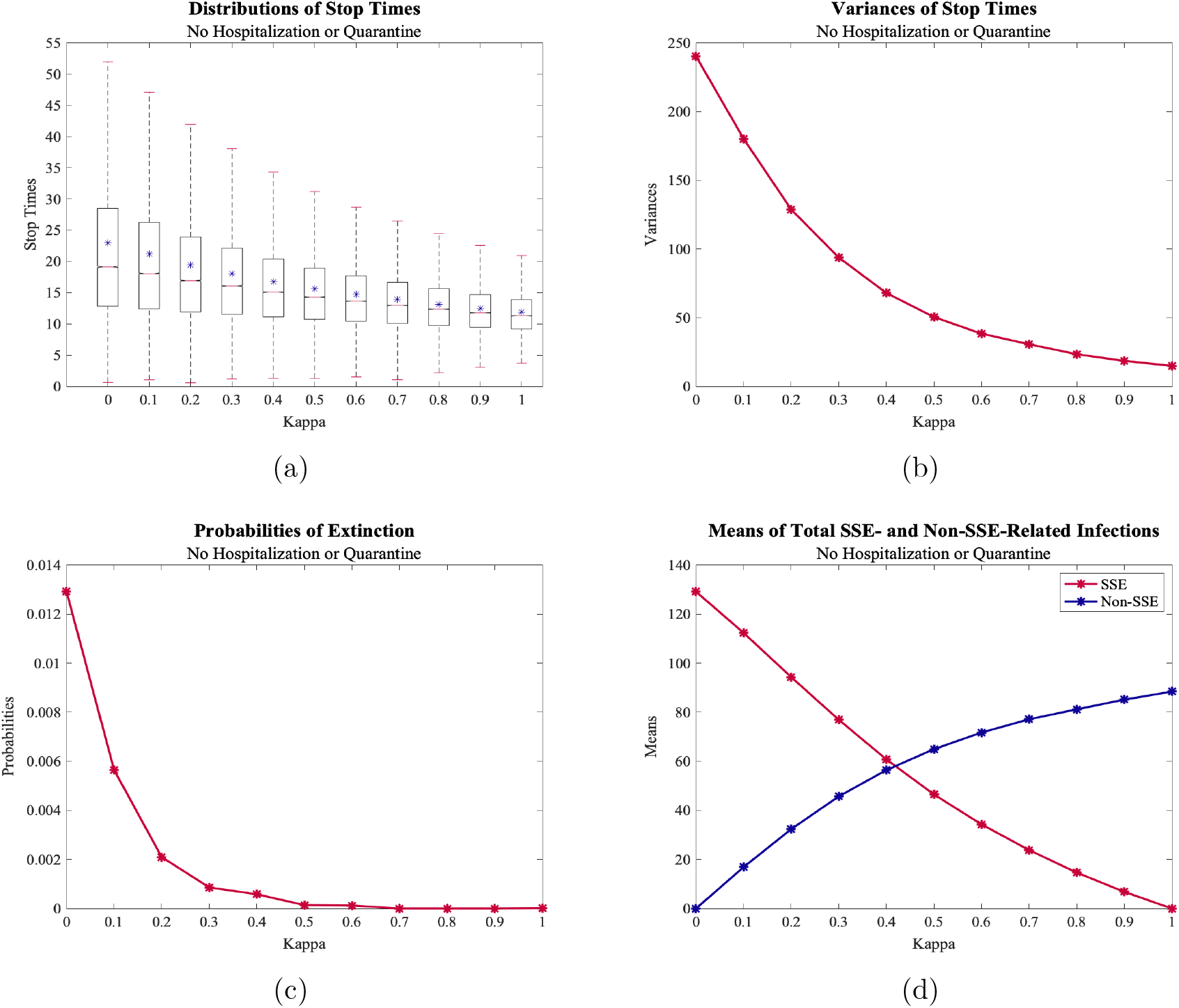
(a) Distributions of Stops Times (NHQ) (b) Variances of Stop Times for NHQ (c) Probabilities of Extinction for NHQ (d) Means of Cumulative Total of SSE- and Non-SSE-Related Infections for NHQ

### Scenario (ii): Realistic Hospitalization and Quarantine (RHQ)

In RHQ, hospitalization, quarantine, premature hospital discharge, and quarantine violation are included in the model. Figure 3(a) shows the distribution of stop times. The maximum, mean, and median stop times strictly decrease for *κ* ∈ [0, 1], while the minimum stop times are similar for *κ* ∈ [0, 0.5) but generally increase for *κ* ∈ [0.5, 1]. Figures 3(b) and 3(c) show the variances of stop times and probabilities of extinction, respectively. Both strictly decrease for *κ* ∈ [0, 1], but they do so more quickly for smaller *κ*, the exception being the variances of stop times for *κ* ∈ [0, 0.1]. Figure 3(d) shows the means of the cumulative numbers of SSE-related and non-SSE-related infections. Their curves are labelled as SSE and Non-SSE in the legend. The SSE curve strictly decreases with increasing *κ*, while the Non-SSE curved stricly increases. They do so more quickly for smaller *κ* and intersect between *κ* = 0.4 and *κ* = 0.5.

**Figure 3:**
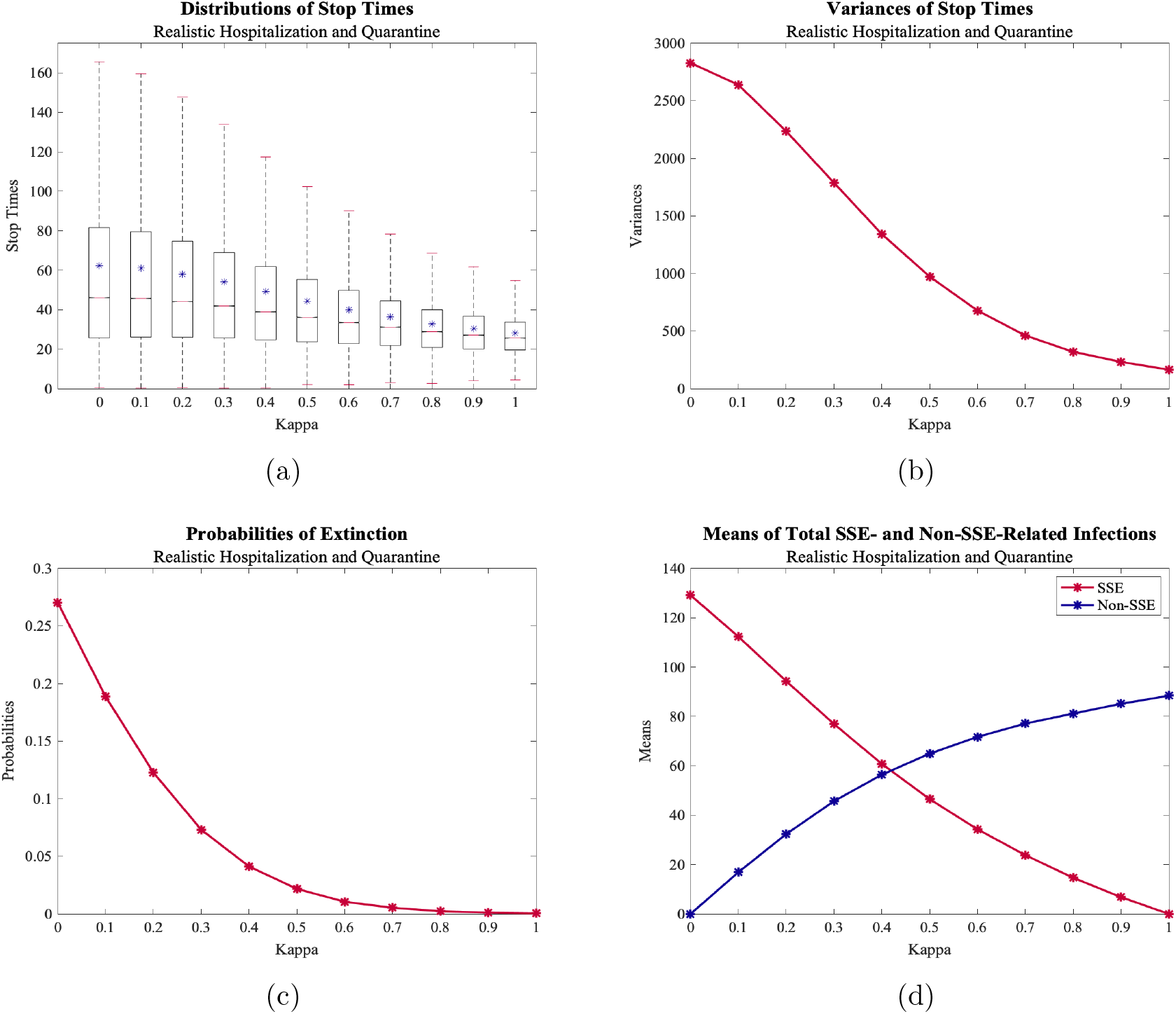
(a) Distributions of Stops Times (RHQ) (b) Variances of Stop Times for RHQ (c) Probabilities of Extinction for RHQ (d) Means of Cumulative Total of SSE- and Non-SSE-Related Infections for RHQ

### Scenario (iii): Idealistic Quarantine and Hospitalization (IHQ)

In IHQ, hospitalization and quarantine are included in the model, but premature hospital discharge and quarantine violation are excluded. Figure 4(a) shows the distribution of stop times. The maximum, mean, and median stop times strictly increase for *κ* ∈ [0, 0.7) but strictly decrease for *κ* ∈ [0.7, 1]. Meanwhile, the minimum stop times are similar for *κ* ∈ [0, 0.8) but strictly increase for *κ* ∈ [0.8, 1]. Figure 4(b) shows the variances of stop times, which strictly increase for *κ* ∈ [0, 0.7) but strictly decrease for *κ* ∈ [0.7, 1]. Figure 4(c) shows the probabilities of extinction, which strictly decrease with increasing *κ* for *κ* ∈ [0, 1] but remain near 1 for *κ* ∈ [0, 0.5) and quickly decrease for *κ* ∈ [0.5, 1]. Figure 4(d) shows the means of the cumulative numbers of SSE-related and non-SSE-related infections. Their curves are labelled as SSE and Non-SSE in the legend. The SSE curve remains approximately constant for *κ* ∈ [0, 0.5) but strictly decreases for *κ* ∈ [0.5, 1]. Meanwhile, the Non-SSE curve strictly increases for *κ* ∈ [0, 0.7), remains approximately constant for *κ* ∈ [0.7, 0.8), and strictly decreases for *κ* ∈ [0.8, 1]. The curves intersect between *κ* = 0.4 and *κ* = 0.5.

**Figure 4:**
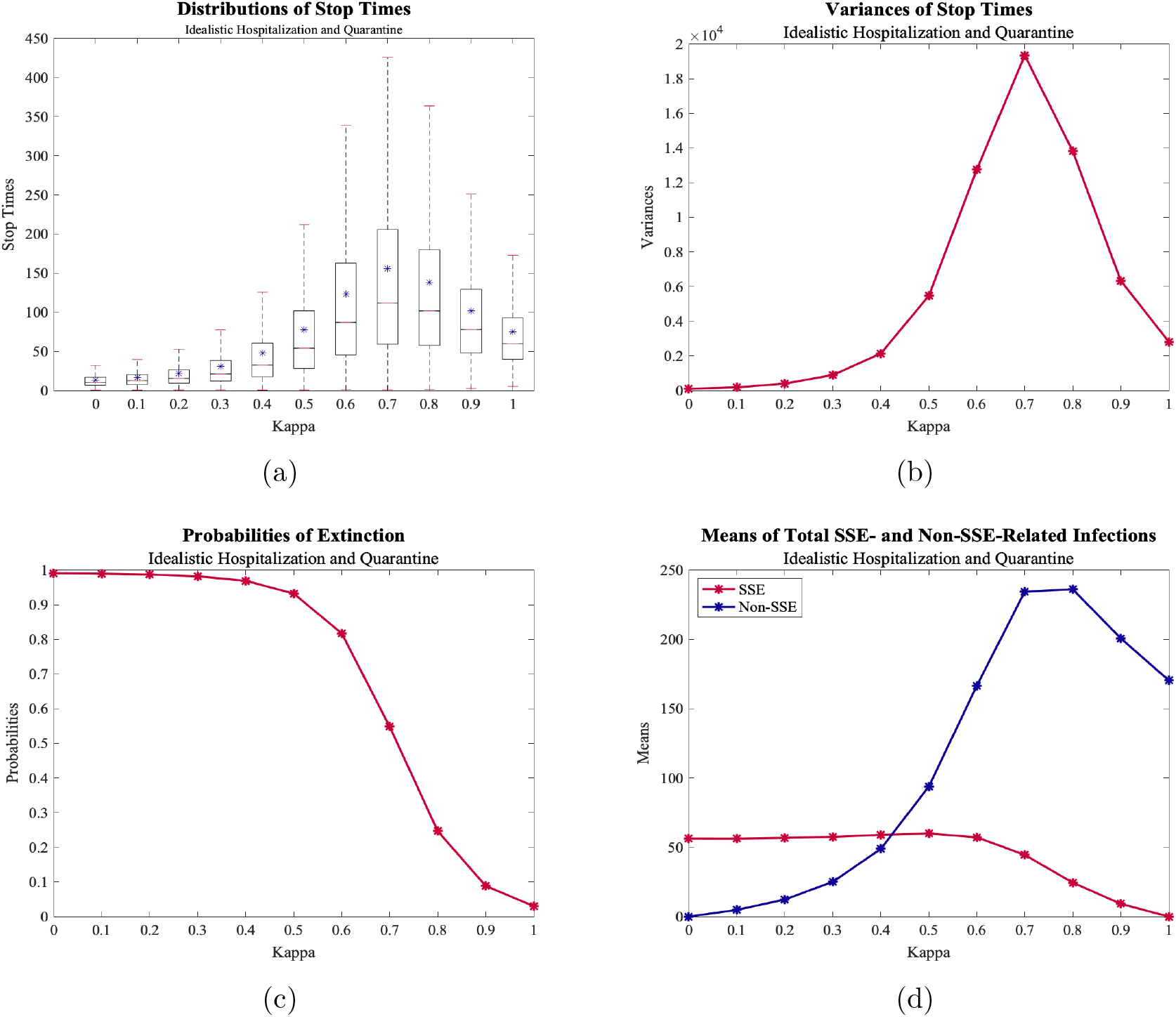
(a) Distributions of Stops Times (b) Variances of Stop Times for IHQ (c) Probabilities of Extinction for IHQ (d) Means of Cumulative Total of SSE- and Non-SSE-Related Infections for IHQ

### Varying Quarantine Violation (RHQ_*lqv*_ and RHQ_*hqv*_)

In RHQ_*lqv*_, RHQ’s quarantine violation level is halved, and in RHQ_*hqv*_, RHQ’s quarantine violation level is doubled. Figures 5(a), 5(b), 5(c), and 5(d) contain curves for RHQ_*lqv*_, RHQ, and RHQ_*hqv*_; these curves are labelled as Low Quarantine Violation, Medium Quarantine Violation, and High Quarantine Violation in the legends. Figures 5(a) and 5(b) show the means and variances of stop times for each quarantine violation level. For RHQ_*lqv*_, both strictly increase for *κ* ∈ [0, 0.2) but strictly decrease for *κ* ∈ [0.2, 1]; for RHQ and RHQ_*hqv*_, both strictly increase for *κ* ∈ [0, 1]. The means and variances of stop times strictly decrease going from RHQ_*lqv*_ to RHQ to RHQ_*hqv*_ for each *κ* ∈ [0, 1], but their differences are greater for smaller *κ*. Figure 5(c) shows the probabilities of extinction. They strictly decrease for *κ* ∈ [0, 1] but do so more quickly for smaller *κ*. The probabilities of extinction also strictly decrease going from RHQ_*lqv*_ to RHQ to RHQ_*hqv*_ for *κ* ∈ [0, 1], but their differences are greater for smaller *κ*. They decrease more quickly for RHQ_*lqv*_ than RHQ and RHQ_*hqv*_. Figure 5(d) shows the cumulative numbers of SSE-related infections. They strictly decrease for *κ* ∈ [0, 1] but do so more quickly for smaller *κ*, the exception being RHQ_*lqv*_ for *κ* ∈ [0, 0.2).

**Figure 5:**
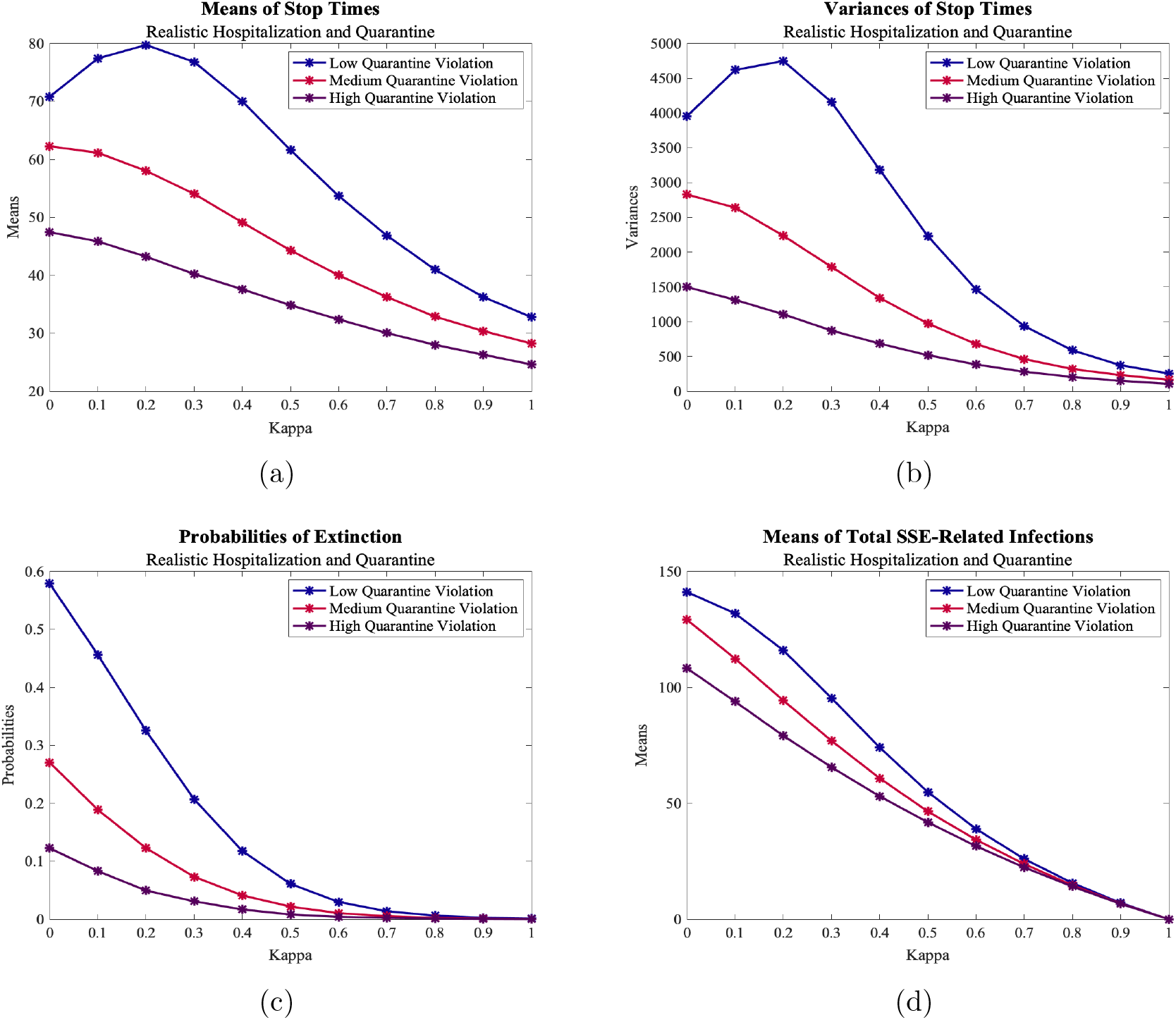
(a) Means of Stops Times for RHQ_*lqv*_, RHQ, and RHQ_*hqv*_ (b) Variances of Stop Times for RHQ_*lqv*_, RHQ, and RHQ_*hqv*_ (c) Probabilities of Extinction for RHQ_*lqv*_, RHQ, and RHQ_*hqv*_ (d) Means of Cumulative Total of SSE-Related Infections for RHQ_*lqv*_, RHQ, and RHQ_*hqv*_

## 4 Discussion

In what follows, recall that as *κ* increases, SSE rates decrease, and as SSE rates decrease, non-SSE rates increase. The relative SSE and non-SSE rates are governed by the constancy condition in Section 2.3. For both NHQ and RHQ, most outbreaks are more variable but less severe when SSE rates are higher, as evidenced by the variances of stop times in Figures 2(b) and 3(b) and the distributions of stop times in Figures 2(a) and 3(a). Here, severity is based on stop times; more severe outbreaks are quicker to attain 50 active infections, while less severe outbreaks are slower. Though most SSE-dominated outbreaks are less severe, the most severe SSE-dominated outbreaks are more severe than the most severe non-SSE-dominated outbreaks, as evidenced by the minimum stop times in Figures 2(a) and 3(a). Outbreaks are also more likely to go extinct when SSE rates are higher, as evidenced by the probabilities of extinction in 2(c) and 3(c). All of the aforementioned observations hold for IHQ when *κ* ∈ [0.7, 1] but not when *κ* ∈ [0, 0.7). For IHQ when *κ* ∈ [0, 0.7), most outbreaks are less variable (see Figure 4(b)) but more severe (see Figure 4(a)) when SSE rates are higher. The most severe SSE-dominated outbreaks are still more severe than the most severe non-SSE-dominated outbreaks (see Figure 4(a)), and outbreaks are still more likely to go extinct (see Figure 4(c)) when SSE rates are higher. Going from NHQ (*κ* ∈ [0, 1]) to RHQ (*κ* ∈ [0, 1]) to IHQ (*κ* ∈ [0.7, 1]), outbreaks increase in variability (see Figures 2(b), 3(b), and 4(b)), decrease in severity (see Figures 2(a), 3(a), and 4(a)), and are more likely to go extinct (see Figures 2(c), 3(c), and 4(c)).

Differences for IHQ when *κ* ∈ [0, 0.7) are likely related to high probabilities of extinction. For *κ* ∈ [0, 0.7), outbreaks may be less variable but more severe when SSE rates are higher because surviving outbreaks are restricted to those with many SSE-related infections early on. This is evidenced by the means of cumulative SSE-related infections remaining approximately constant when *κ* ∈ [0, 0.5) despite decreasing SSE rates, after which they decrease with decreasing SSE rates, as expected (see Figure 4(d)). Meanwhile, the means of total non-SSE-related infections increase with increasing non-SSE rates when *κ* ∈ [0, 0.7), as expected, but decrease when *κ* ∈ [0.7, 1] despite increasing non-SSE rates (see Figure 4(d)). The weakening pull towards extinction may allow 50 current infections to be attained more quickly (see Figure 4(a)), to the extent that the number of cumulative infections is reduced. For NQH and RHQ, the probabilities of extinction are lower than for IHQ (see Figures 2(c), 3(c), and 4(c)), and the means of cumulative SSE-related and non-SSE-related infections for NHQ and RHQ decrease with decreasing SSE rates and increase with increasing non-SSE rates (see Figures 2(d) and 3(d)), as expected. Note that for NHQ and RHQ, the means of cumulative SSE-related infections decrease more quickly for smaller *κ* (see Figures 2(d) and 3(d)), while for IHQ, the means of cumulative SSE-related infections decrease more slowly for smaller *κ* when *κ* ∈ [0.5, 0.7) but more quickly for smaller *κ* when *κ* ∈ [0.7, 1] (see Figure 4(d)). This, in conjunction with the slight lag between the means of cumulative SSE-related infections decreasing and outbreaks becoming more variable but less severe when SSE rates are higher, suggests that *κ* ∈ [0.5, 0.7) corresponds to a transitional period. During this period, dynamics change from those only observed for IHQ to those also observed for NHQ and RHQ.

Greater variability in SSE-dominated outbreak profiles may limit predictions. Increased variability increases prediction uncertainty, which may decrease prediction accuracy. Increased prediction uncertainty also shortens the time period over which predictions hold. Such limitations impede public health responses. Meanwhile, taking more time in most cases to attain 50 current infections when SSE rates are higher may allow the virus avoid detection for longer; more sensitive and stable surveillance systems are necessary to detect the virus at low levels. Because even slow SSE-dominated outbreaks may abruptly take off, timeliness is also key. For further information on the stability, sensitivity, and timeliness of surveillance systems, see [12, 15]. Persistence at low levels also facilitates the evolution of more infectious variants [5, 46]. The public health consequences associated with SSE-dominated outbreaks are, in some cases, mitigated by high probabilities of extinction. For IHQ, SSE-dominated outbreaks almost always go extinct; for RHQ, SSE-dominated outbreaks are somewhat likely to go extinct. Public health consequences are thus mostly mitigated for IHQ and somewhat mitigated for RHQ. They are not, however, mitigated for NHQ, as outbreaks (SSE- or non-SSE-dominated) seldom go extinct in this scenario.

Across-the-board decreases in probabilities of extinction going from IHQ to RHQ to NHQ suggest that increasing hospitalisation and quarantine and decreasing premature hospital discharge and quarantine violation forces more outbreaks to extinction. Greater differences for SSE-dominated outbreaks versus non-SSE-dominated outbreaks imply that hospitalisation and quarantine are more effective at controlling SSE-dominated outbreaks; likewise, premature hospital discharge and quarantine violation are more consequential for such outbreaks. Quicker decreases in probabilities of extinction for IHQ, RHQ, and NHQ when SSE rates are higher further imply that SSE-dominated outbreaks are more sensitive to changes in hospitalisation, quarantine, premature hospital discharge, and quarantine violation. High probabilities of extinction for IHQ when SSE rates are higher, in conjunction with the quicker decreases, imply that reducing premature hospital discharge and quarantine violation substantially increases the effectiveness of hospitalisation and quarantine at forcing SSE-dominated outbreaks to extinction. When weighing the benefits and detriments of control measures, the relative rates of SSEs versus non-SSEs should thus be considered; for more on the benefits and detriments of control measures, see [27, Chapter 17]. Note that in all of the aforementioned, the effectiveness of hospitalisation and quarantine and the consequences of premature hospital discharge and quarantine violation were evaluated using probabilities of extinction versus other metrics, such as cumulative infections or deaths; the latter metrics would be misleading because simulations were ended once either the disease went extinct or 50 active infections were attained (see Section 2).

For RHQ_*lqv*_, outbreaks are more variable but less severe on average when SSE rates are higher (see Figures 5(b) and 5(a)), similar to NHQ and RHQ; for RHQ_*hqv*_, outbreaks are less variable but more severe on average for *κ* ∈ [0, 0.2) and more variable but less severe on average for *κ* ∈ [0.2, 1] when SSE rates are higher (see Figures 5(b) and 5(a)), similar to IHQ. For RHQ_*lqv*_ and RHQ_*hqv*_, outbreaks are more likely to go extinct when SSE rates are higher (see Figure 5(c)), similar to NHQ, RHQ, and IHQ. The means of total SSE-related infections decrease with decreasing SSE rates for RHQ_*lqv*_, RHQ, and RHQ_*hqv*_, but they do so more slowly for RHQ_*lqv*_ than RHQ or RHQ_*hqv*_ when SSE rates are higher (see Figure 5(d)). We expect the opposite, given that quarantined individuals cannot cause SSEs. Going from RHQ to RHQ_*hqv*_, the means of total SSE-related infections decrease more slowly, as expected. Differences for RHQ_*lqv*_ when *κ* ∈ [0, 0.2) may thus be related to high probabilities of extinction (see Figure 5(c)), with *κ* ∈ [0, 0.2) corresponding to a transitional period, similar to IHQ with *κ* ∈ [0.5, 0.7).

Going from RHQ_*lqv*_ (*κ* ∈ [0, 1]) to RHQ (*κ* ∈ [0, 1]) to RHQ_*hqv*_ (*κ* ∈ [0.2, 1]), outbreaks decrease in variability, increase in severity, and are less likely to go extinct (see Figures 5(b), 5(a), and 5(c)). These differences are greater for SSE-dominated outbreaks, most notably with respect to variability and extinction. Taking less time on average to attain 50 current infections when quarantine violation is higher shortens the time period during which outbreaks may feasibly be contained; the further outbreaks progress, the harder it becomes to contain them [33]. While lesser variability in outbreak profiles when quarantine violation is higher allows for lesser prediction uncertainty, it also allows for a greater number of more severe outbreaks, given that outbreaks are more severe on average when quarantine violation is higher. Larger numbers of infections over shorter time periods may strain resources [14]. Across-the-board decreases in probabilities of extinction going from RHQ_*lqv*_ to RHQ to RHQ_*hqv*_ suggest that decreasing quarantine violation forces more outbreaks to extinction. Greater differences for SSE-dominated versus non-SSE-dominated outbreaks imply that quarantine violation is more consequential for SSE-dominated outbreaks. Quicker decreases in probabilities of extinction for RHQ_*lqv*_, RHQ, and RHQ_*hqv*_ when SSE rates are higher further imply that SSE-dominated outbreaks are more sensitive to changes in quarantine violation. High probabilities of extinction for RHQ_*lqv*_ when SSE rates are higher, in conjunction with the quicker decreases, imply that reducing quarantine violation substantially increases the effectiveness of quarantine at forcing SSE-dominated outbreaks to extinction. This reinforces our previous findings regarding premature hospital discharge and quarantine violation. Again note that in all of the aforementioned, the effectiveness of quarantine and consequences of quarantine violation were evaluated using probabilities of extinction versus other metrics, such as cumulative infections or deaths; the latter metrics would be misleading because simulations were ended once either the disease went extinct or 50 active infections were attained (see Section 2).

### Comparison with SI models in literature

Lloyd, et al [32] and James, et al [26] utilize discrete-time Markov Chain (DTMC) models to investigate the relative influences of SIs and SSEs on outbreak dynamics. Their models’ underlying biological process is more simplistic than ours; susceptible individuals are infected and infected individuals recover. There are neither exposed, asymptomatic, hospitalized, nor quarantined individuals. DTMC models also evolve through time differently, and their state changes have different meanings (see Section 2.5).

Lloyd, et al [32] and James, et al [26] found that increasing SIs or SSEs relative to non-SIs or non-SSEs increases the probability that outbreaks go extinct but decreases the variability of surviving outbreaks, which are more severe when dominated by SIs or SSEs. This partialy agrees with our findings for NHQ (*κ* ∈ [0, 1]), RHQ (*κ* ∈ [0, 1]), and IHQ (*κ* ∈ [0.7, 1]) – increasing SSEs relative to non-SSEs increases both the probability that outbreaks go extinct and the variability of surviving outbreaks, which are less severe when dominated by SSEs – and fully agrees with our findings for IHQ (*κ* ∈ [0, 0.7)). While the probabilities of extinction increase with increasing influence of SIs or SSEs relative to non-SIs or non-SSEs for Lloyd et al’s, James et al’s, and our model, they approach different values. For the individual-based model, they approach 1, but for the event-based models, they do not. This suggests that SSE-dominated outbreaks are more likely to survive than SI-dominated outbreaks.

Differences in the variability and severity of surviving outbreaks between Lloyd, et al’s [32], James et al’s [26], and our model for NQH (*κ* ∈ [0, 1]), RHQ (*κ* ∈ [0, 1]), and IHQ (*κ* ∈ [0, 0.7)) may be related to differences in probabilities of extinction, which are higher for Lloyd, et al’s [32], James, et al’s [26], and our model for IHQ (*κ* ∈ [0.7, 1]). As discussed above, high probabilities of extinction may limit surviving outbreaks to more severe outbreaks. As the pull towards extinction weakens, a larger variety of outbreaks may survive. Recall that, while most SSE-dominated outbreaks are less severe than non-SSE-dominated oubreaks for NHQ (*κ* ∈ [0, 1]), RHQ (*κ* ∈ [0, 1]), and IHQ (*κ* ∈ [0.7, 1]), the most severe SSE-dominated outbreaks are more severe than the most severe non-SSE-dominated outbreaks; this implies that, if sufficiently many of the less severe SSE-dominated outbreaks were to go extinct, most of the surviving SSE-dominated outbreaks may be more severe than the non-SSE-dominated outbreaks, which would agree with the Lloyd, et al’s, James, et al’s, and our results for IHQ (*κ* ∈ [0, 0.7)). Overall, differences between the individual-based and event-based models suggest that, if outbreaks are SI- and/or SSE-dominated, SIs and SSEs must be considered separately, as they have distinct influences on outbreak dynamics.

## 5 Conclusion

This study had three main goals: investigating (1) the influence of SSEs relative to that of non-SSEs on outbreak dynamics, (2) the effectiveness of hospitalisation and quarantine as control measures for SSE- versus non-SSE-dominated outbreaks, and (3) the influence of quarantine violation on the effectiveness of quarantine for SSE- versus non-SSE-dominated outbreaks.

To accomplish these goals, we incorporated SSEs into a continuous-time Markov chain (CTMC) model (Section 2.2), imposed a constancy condition (Section 2.3), simulated the CTMC model under multiple scenarios (Section 2.4), and varied quarantine violation levels (Section 2.4). The scenarios differed in their inclusion/exclusion of hospitalisation, quarantine, premature hospital discharge, and quarantine violation.

When hospitalisation and quarantine are excluded or when hospitalisation, quarantine, premature hospital discharge, and quarantine violation are included, SSE-dominated outbreaks are more variable, less severe, and more likely to go extinct than nonSSE-dominated outbreaks. While most SSE-dominated outbreaks are less severe, the most severe SSE-dominated outbreak is more severe than the most severe nonSSE-dominated outbreaks. When hospitalisation and quarantine are included but premature hospital discharge and quarantine violation are excluded, SSE-dominated outbreaks are more likely to go extinct than non-SSE-dominated outbreaks but less variable and more severe. When hospitalisation, quarantine, premature hospital discharge, and quarantine violation are included and quarantine violation is halved, SSE- dominated outbreaks behave similarly to when hospitalisation and quarantine are included but premature hospital discharge and quarantine violation are excluded; when quarantine violation is doubled, outbreaks behave similarly to when hospitalisation and quarantine are excluded.

In all scenarios, hospitalisation and quarantine are more effective at controlling SSE- dominated outbreaks than non-SSE-dominated outbreaks. Similarly, premature hospital discharge and quarantine violation are more consequential for SSE-dominated outbreaks. When hospitalisation and quarantine are excluded, SSE-dominated outbreaks are highly unlikely to go extinct; when hospitalisation, quarantine, premature hospital discharge, and quarantine violation are included, SSE-dominated outbreaks are moderately unlikely to go extinct; and when hospitalisation and quarantine are included but premature hospital discharge and quarantine violation are excluded, SSE- dominated outbreaks are highly likely to go extinct. Halving quarantine violation significantly increases the likelihood that SSE-dominated outbreaks go extinct, while doubling quarantine violation somewhat decreases the likelihood that SSE-dominated outbreaks go extinct.

Altogether, SSE-dominated outbreaks notably differ from non-SSE-dominated outbreaks in terms of variability, severity, and likelihood of extinction; they also differ from SI-dominated outbreaks, albeit more subtly (see Section 4). SSE-dominated outbreaks’ dynamics are strongly influenced by whether individuals may be hospitalised or quarantined, as well as whether they may be prematurely discharged from the hospital or violate quarantine. Both hospitalisation and quarantine are highly effective control measures for SSE-dominated outbreaks, but premature hospital discharge and quarantine violation substantially reduce their effectiveness. Note that we evaluated control measures using the likelihood of extinction. All of the aforementioned has important public health implications (see Section 4), which necessitates that SARS-CoV-2 modelers 1) determine the extent to which SSEs and/or SIs contribute to spread and 2) distinguish between SSEs, SIs, and non-SSEs/non-SIs in their models. Finally, further exploration of the individual and combined influences of SSEs and SIs on outbreak dynamics, as well as the effectiveness of control measures for different types of outbreaks, is necessary to better inform containment and eradication efforts.

## Data Availability

All data produced in the present work are contained in the manuscript

## Data Availability

All data used in this study came from published, cited sources, and are included in the text.

## Conflicts of Interest

The authors declare that there are no conflicts of interest.

## Author contributions

Jordan Bramble: Formal analysis, visualization, software, writing - original draft preparation, reviewing, and editing; Alexander Fulk: Conceptualization, formal analysis, writing - reviewing and editing; Raul Saenz: Conceptualization, writing - review, and editing; Folashade B. Agusto: Conceptualization, methodology, project administration, supervision, writing- reviewing and editing, funding acquisition.

## Acknowledgement

This research is supported by National Science Foundation under the grant number DMS 2028297.

## Notes

### Competing Interest Statement

The authors have declared no competing interest.

### Author Declarations

The study used openly available data that were originally used in Agusto et al. "To isolate or not to isolate: The impact of changing behavior on COVID-19 transmission." BMC Public Health 22.1 (2022): 1-20.

